# COVID-19: Easing the coronavirus lockdowns with caution

**DOI:** 10.1101/2020.05.10.20097295

**Authors:** Akpojoto Siemuri, Rasheed Omobolaji Alabi, Mohammed Elmusrati

**Affiliations:** Department of Industrial Digitalization, School of Technology and Innovations, University of Vaasa, Vaasa, Finland

**Author notes:** **The last two authors have equal contributions and first authorship**. **Corresponding Author: Rasheed Omobolaji Alabi**, Department of Industrial Digitalization, School of Technology and Innovations, University of Vaasa, Vaasa, Finland. **E-mail address:**.

**Keywords:** COVID, Lockdowns, Ease, Forecasting, SARS-COV-2

## Abstract

**Background:** The spread of the novel severe acute respiratory syndrome coronavirus (SARS-CoV-2) has reached a global level, creating a pandemic. The government of various countries, their citizens, politicians, and business owners are worried about the unavoidable economic impacts of this pandemic. Therefore, there is an eagerness for the pandemic peaking.

**Objectives:** This study uses an objective approach to emphasize the need to be pragmatic with easing of lockdowns measures worldwide through the forecast of the possible trend of COVID-19. This is necessary to ensure that the enthusiasm about SARS-CoV-2 peaking is properly examined, easing of lockdown is done systematically to avoid second-wave of the pandemic.

**Methods:** We used the Facebook prophet on the World Health Organization data for COVID-19 to forecast the spread of SARS-CoV-2 for the 7^th^ April until 3rd May 2020. The forecast model was further used to forecast the trend of the virus for the 8^th^ until 14^th^ May 2020. We presented the forecast of the confirmed and death cases.

**Results:** Our findings from the forecast showed an increase in the number of new cases for this period. Therefore, the need for easing the lockdown with caution becomes imperative. Our model showed good performance when compared to the official report from the World Health Organization. The average forecasting accuracy of our model was 79.6%.

**Conclusion:** Although, the global and economic impact of COVID-19 is daunting. However, excessive optimism about easing the lockdown should be appropriately weighed against the risk of underestimating its spread. As seen globally, the risks appeared far from being symmetric. Therefore, the forecasting provided in this study offers an insight into the spread of the virus for effective planning and decision-making in terms of easing the lockdowns in various countries.

## 1. Introduction

The year 2019 ended on a remarkable note over the novel severe acute respiratory syndrome coronavirus (SARS-CoV-2) [1]. Patients with SARS-CoV-2 infection can develop coronavirus disease 2019 (COVID-19) [2] with an incubation period ranging from 2 to 14 days [1]. The first case of the coronavirus disease 2019 (COVID-19) was reported in Wuhan, Hubei province, China in December 2019 [3]. It was reported to have originated in bats and transmitted to humans through yet unknown animals within Wuhan, China. From the primary place where it first emerged, it has spread globally. Thus, the World Health Organization (WHO) declared the outbreak a pandemic [4].

The mode of transmission in humans includes inhalation or contact with infected droplets where the symptoms include fever, cough, sore throats, difficulty in breathing properly, fatigue among others [1]. Of note, many people are asymptomatic (showing no symptoms of the virus), mild symptoms in some people, and severe symptoms in others especially the elderly and those with comorbidities. These severe symptoms may progress to multi-organ dysfunction, acute respiratory distress syndrome, respiratory failure, and pneumonia [1]. The severe symptoms have usually resulted in high rates of contacts to the healthcare providers, hospitalization, and admission to the intensive care units (ICU) [5]

Around 96, 000 cases were reported in early March 2020 (05/03/2020) [1]. By early April, the number of cases globally has increased to 1,603,330 (10/03/2020) [6]. The rise in the number of cases had resulted in various measures to reduce the spread of this virus. The example of such measures includes lockdown by countries [7]. Other measures include social distancing, contact tracing, quarantine, increased handwashing and sanitizing, temporary closedown of schools, businesses, and international trades. Therefore, the global economic impact of this pandemic is of great concern.

Entrepreneurs have been rendered temporarily unemployed. Governments have either introduced laws, update social welfare, or provides palliatives to ensure that the overall impacts of this pandemic on her citizens are relatively reduced. Though, the long term sustainability of these approaches remains doubtful. Thus, Government, politicians, and business owners are eager to get back to their normal daily routine. This eagerness has led to optimism about the pandemic peaking.

Despite the significant economic impacts of the COVID-19 pandemic for the past 100 days (since the virus had been identified), reasonable caution against excessive optimism about the pandemic peaking must be exercised to minimize the overall impacts (economic, social, and death) [8]. Therefore, this paper introduces an objective approach to forecasting the global spread of COVID-19 crisis. Accurate forecasting of the associated risks and spread of the virus is necessary to ensure that the implications can be properly analyzed, planning by the government and business owners improved, and informed decisions about the pandemic can be made in terms of easing the lockdown.

## 2. Methods

### 2.1. World Health Organization (WHO) data

The study data was obtained from the World Health Organization (WHO). The WHO COVID-19 data was used to assess the worldwide and country-specific trends of COVID-19 new confirmed and death cases around the world. The data contained cases from 09 January 2020 until 6 April 2020. The input variables contained in the dataset were day, country, country name, region, death, cumulative deaths, confirmed, cumulative confirmed. As the data is publicly available, informed consent was not needed.

### 2.2. Trend of the virus

The forecasting of the trend of the virus was done using Facebook’s Prophet in Google Colaboratory (a free Jupyter notebook environment). The dataset and the corresponding libraries were imported to Jupyter Notebook. The data was further preprocessed to remove missing values and ensure that the data is in the proper format. Additionally, the columns were renamed for convenience in the analyses. The visualization of the top 11 countries, Finland, and Nigeria (most populous African country) as of 7^th^ May 2020 is given in Figure 1. The actual decomposition of the time series data was done using an additive model. The additive model has been implemented because it is easy to develop, fast to train, and provides interpretable patterns ^9^ We used the last 3 months of the COVID-19 cases to forecast for the next 5 days (8^th^ until 14^th^ May 2020) using the Facebook Prophet, with a 95% prediction interval by creating a base model using seasonality-related parameters and additional regressors. Note that the yearly and daily seasonality was not set since COVID-19 is not a seasonal disease. The forecast given by our model was compared with the official data reported on the World Health Organization (WHO) official website for COVID-19 updates for the same date. The dashboard of WHO’s official website for comparison is available at https://covid19.who.int/. The result of the comparison for 07.04.2020 – 03.05.2020 is presented in Table 1. We also presented the model forecast for the 8^th^ until 14^th^ May 2020 to further validate our model prior to the official release of the cases by the World Health Organization.

**Figure 1.**
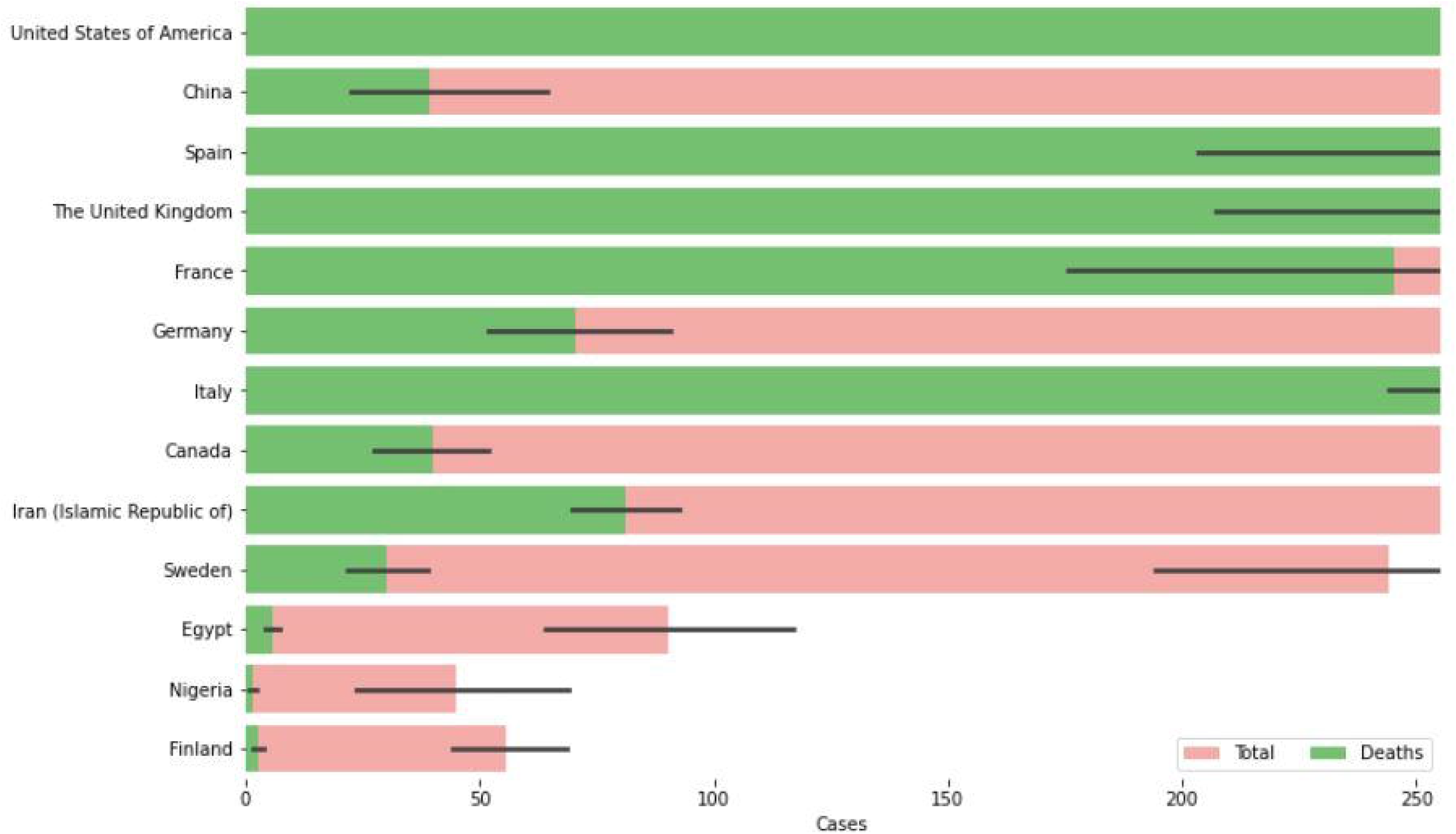
The number of confirmed cases and death in top 11 countries in the world, Finland and Nigeria.

**Table 1.** The comparison of the forecast model with the reported cases on WHO’s official website.

| Validated Forecasted data |  |  |  |  |  |  |
| --- | --- | --- | --- | --- | --- | --- |
| Date | Model Forecast for confirmed cases | WHO Reported cases for confirmed cases | Model Accuracy | Model Forecast for death cases | WHO Reported cases for death cases | Model Accuracy |
| 7.4.2020 | 84833 | 68465 | 76,1 | 5623 | 5009 | 87,7 |
| 8.4.2020 | 88895 | 73849 | 79,6 | 5975 | 6685 | 89,4 |
| 9.4.2020 | 93295 | 82736 | 87,2 | 6278 | 6325 | 99,3 |
| 10.4.2020 | 95575 | 84635 | 87,1 | 6467 | 7231 | 89,4 |
| 11.4.2020 | 100173 | 90778 | 89,7 | 6891 | 6946 | 99,2 |
| 15.4.2020 | 92791 | 69728 | 66,9 | 7154 | 5909 | 78,9 |
| 16.4.2020 | 98110 | 77846 | 74,0 | 7394 | 7911 | 93,5 |
| 17.4.2020 | 86878 | 82622 | 94,8 | 7597 | 8473 | 89,7 |
| 18.4.2020 | 88480 | 85501 | 96,5 | 6926 | 6681 | 96,3 |
| 19.4.2020 | 87380 | 82180 | 93,7 | 6860 | 6507 | 94,6 |
| 20.4.2020 | 90228 | 72397 | 75,4 | 6950 | 5261 | 67,9 |
| 21.4.2020 | 91276 | 83596 | 90,8 | 7083 | 5118 | 61,6 |
| 22.4.2020 | 92340 | 73448 | 74,3 | 7430 | 6060 | 77,4 |
| 23.4.2020 | 97801 | 73911 | 67,7 | 7747 | 6675 | 83,9 |
| 24.4.2020 | 95541 | 82089 | 83,6 | 7562 | 6290 | 79,8 |
| 28.4.2020 | 94744 | 73759 | 71,5 | 6925 | 3894 | 22,2 |
| 29.4.2020 | 95283 | 66321 | 56,3 | 7297 | 5378 | 64,3 |
| 30.4.2020 | 99878 | 72955 | 63,1 | 7598 | 9785 | 77,6 |
| 1.5.2020 | 101002 | 84983 | 81,2 | 7719 | 6405 | 79,5 |
| 2.5.2020 | 104273 | 90520 | 84,8 | 7808 | 5803 | 65,4 |
| 3.5.2020 | 102431 | 83985 | 78,0 | 7721 | 8626 | 89,5 |
|  |  | Average | 79,6 |  | Average | 80,3 |

## 3. Results

The study cohort included 210 countries at the time of data extraction from WHO official website (06.04.2020). After building the forecast model, it showed good forecasting accuracy in predicting the confirmed and death cases for the date of data retrieval (6.04.2020) as shown in Table 1, respectively. As shown in Table 1, the prediction accuracy of the model for the confirmed cases was 79.6% and 80.3% (average of 79.9%) for the death cases when its performance for forecasting for the period 07.04.2020 – 03.05.2020 was compared with the reported cases on WHO official website for confirmed and death cases, respectively. The details of the forecast for this period is given in Table 1. As shown in Figures 2-3, there was increase in the number of cases, although not spike increase in the number of confirmed and death cases as seen in previous months. Additionally, the next 7 days (8^th^ – 14^th^ May 2020) prediction of the pandemic COVID-19 showed a rise in the number of confirmed cases and deaths from COVID-19 globally (Figure 2-3). As shown in Figure 2, few cases were confirmed between Mondays until Wednesdays. Likewise, most deaths occurred between Tuesdays and Thursdays (Figure 3). Towards easing the lockdown, the World Health Organization published six (6) important criteria that includes evidence that the transmission of COVID-19 is controlled, effective mechanism of contract tracing, quarantine, and testing, minimized outbreaks amongst the vulnerable in care homes, continuous adherence to preventive measures (physical distancing, handwashing, etc.), importation risks can be managed, and proper information and engagement of the communities concerning the pandemic [9]. Most countries have published tentative plans to ease the lockdown measures to address the spread of the COVID-19 pandemic. As shown by our predictive model, restarting the economies that had been significantly affected by the sudden shutdowns to curb the devastating effects of COVID-19 this should be done with caution [10]. This is necessary to avoid a possible second-wave of the virus which could have more damaging consequences [10].

**Figure 2.**
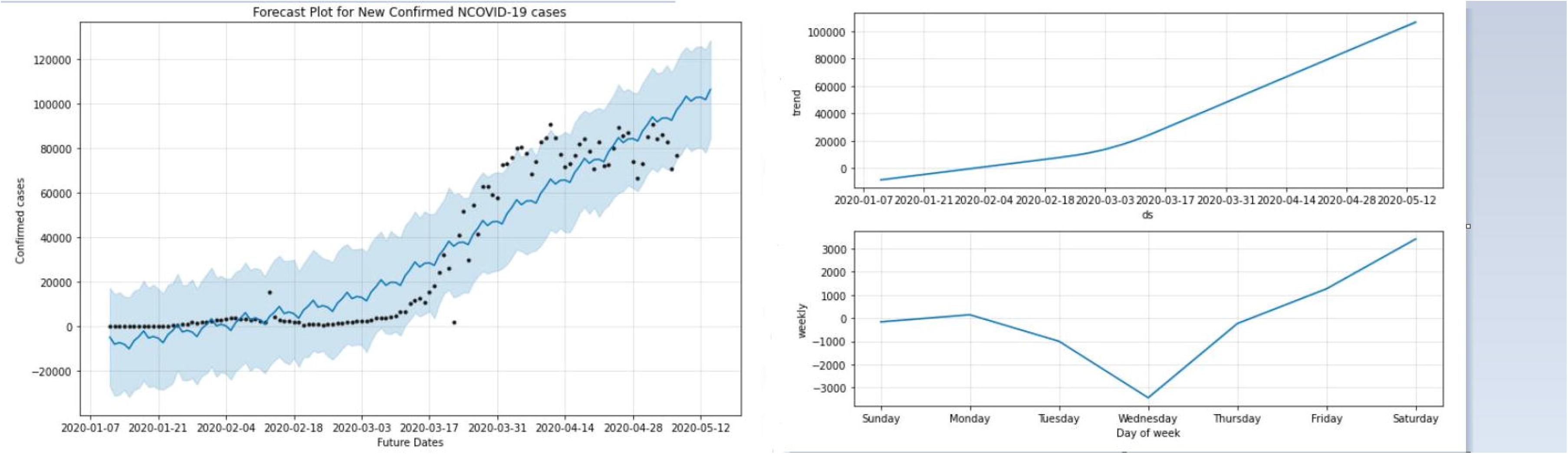
Forecast plot for new confirmed NCOVID-19 cases.

**Figure 3.**
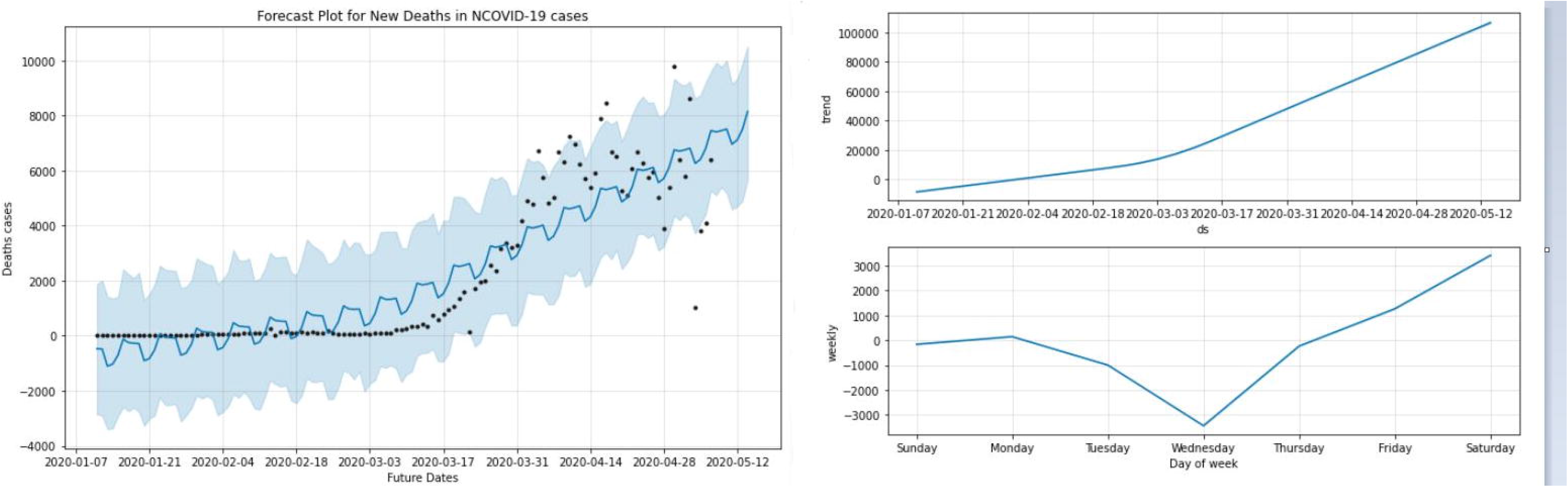
Forecast plot for new death in NCOVID-19 cases.

## 4. Discussion

In this study, we used a time series forecasting approach to forecast the trend of COVID-19 from 07.04.2020 until 03.05.2020 (Table 1) and forecast the trend for the next 7 days (08-14.05.2020). This approach was aimed at emphasizing that the easing of lockdowns in various countries should be done systematically to avoid second-wave of the pandemic. We produced a forecast model using a Facebook prophet that used additive models [11]. A Facebook prophet was used as it is accurate, fast, fully automatic, offers tunable forecast, and compatible with Python or R [12]. Of note, as this is not a seasonal occurrence and the fact that there is no known vaccine yet, we assume that the trend will continue indefinitely in the modelling. The average forecasting accuracy of our model was 79.9%. The differences between our forecast model and the WHO official reported cases may be due to the possible errors, delay or inadequate reporting by various countries. Despite this variation, our forecast model still showed reasonable accuracy to forecast the confirmed and death cases for the target dates. The fact that the confirmed cases increases significantly between Wednesdays and Saturdays may account for the time taken to get and record the results from these tests. Similarly, the curve decreased on the weekend as most death cases were usually reported and recorded during the weekdays.

Our model produced a comparable performance in terms of the trend of the pandemic as shown in Table 1. It was observed that, there was no spike increase in the number of confirmed and death cases within this period. This may suggest that the pandemic may have started to peak in various countries. Despite this, for the period of 8^th^ - 14^th^ May 2002, our model predicted a rise in the number of confirmed cases globally (Figures 2-3). Therefore, excessive optimism about the easing the lockdown in various countries should be met with caution. This is because, at present, the life-and-death pressure on our health facilities is enormous [13-15]. The testing places and hospitals currently have limited or no resources to treat COVID-19 patients [14]. Thus, change of policies to relax the lockdown and social distancing should be done systematically, otherwise, an increase in the number of hospitalized patients may be inevitable. With the current states of the various hospitals globally, it may be difficult to properly manage COVID-19 patients.

The global infection is expected to rise to 40-70% in the coming year [16]. The concern about the rise in the number of confirmed cases was also echoed by the Governor of Massachusetts echoed when he projected that between 0.7 and 2.5 % of the state’s population are likely to contract the coronavirus [17]. The main concern is for how long do we continue to flatten the curve with the lockdown, quarantine, contract tracing, and social distancing measures? Admittedly, the economic and global impact as a result of these measures is massive. Therefore, the concerned authorities (government, politicians and other stakeholders) have started to relax the lockdown policies to minimize the economic impacts.

However, as predicted by our forecast model, a possible rise in the number of confirmed and death cases emphasize the need to have widespread testing in place and hospitals have the resources needed to treat COVID-19 patients prior to switching of gears to allow for controlled exposures [17]. Therefore, the government and policymakers need to get the trend of the spread of the virus right. The understanding of the trajectory of COVID-19 spread ensures that the right decision regarding public policies is made [17]. As such, with the forecast of the possible rise in the number of confirmed and death cases as presented by our model, and the suggestion that only possible COVID-19 vaccine may bring back normalcy [18,19], informing the citizens of the possibility of spending a year or more at home prior to the development of a possible vaccine for COVID-19, or get immune in a hard way becomes pertinent [17].

As pharmaceutical treatments for COVID-19 are yet to be found, the modality of exposure of citizens to the virus needs to be controlled. With herd immunity (exposure to the virus), the vulnerable citizens fall to save the less vulnerable. The main question with this strategy is how this can be done ethically and responsibly. As predicted by our model, the rise in the cases meant that exposure of the world population should be done reasonably, sensibly and controllably for the citizens to acquire herd immunity while the world awaits the breakthrough in the vaccine for COVID-19 [19]. Thus, the government needs to strike a balance between the health and economic impact of this pandemic. If this is not properly considered, there is a high risk of viral reintroduction (those tested negative after initial exposure) [19], intense pressure on health facilities, and increased deaths from COVID-19. As various countries such as France starts to ease the lockdown, important measures such as making face masks compulsory, extra vigilance in the capital regions, reducing the number of public gatherings and events, and temporary closing of place of schools and places worships may still be in effect to avoid a possible second-wave of the pandemic [20].

In conclusion, despite the economic impact of COVID-19, the government and policy-makers in various countries should be very pragmatic in the easing the lockdown measures. Systematic easing of the lockdowns ensures that curve is not only flattened but crushed with controlled exposure to the virus pending the breakthrough of possible pharmaceutical treatment. The fact that our model predicted a spike in the rise should not lead to overly panicking [21] but to understand the trend for the proper decision-making process, especially in high-risk cases. This ensures that we are not only flattening the curve but crushing the curve through controlled herd immunity of the world’s population.

## Data Availability

Public data from World Health Organization about COVID-19

